# Hemodynamic Response after Intra-Aortic Balloon Counter-Pulsation in Cardiac Amyloidosis and Cardiogenic Shock

**DOI:** 10.1101/2023.04.17.23288714

**Authors:** Joshua Longinow, Zachary J. Il’Giovine, Pieter Martens, Andrew Higgins, Lauren Ives, Edward G. Soltesz, Michael Z. Tong, Jerry D. Estep, Randall C. Starling, W.H. Wilson Tang, Mazen Hanna, Ran Lee

## Abstract

**Background:** In those with heart failure-related cardiogenic shock, intra-aortic balloon pump may improve hemodynamics and be useful as a bridge to advanced therapies. We explore whether those with cardiac amyloidosis and heart failure-related cardiogenic shock might experience hemodynamic improvement and describe the hemodynamic response after intra-aortic balloon pump.

**Methods:** We retrospectively identified consecutive patients with a diagnosis of cardiac amyloid, either light-chain or transthyretin, who were admitted to our intensive care unit with heart failure-related cardiogenic shock. Patients were excluded if intra-aortic balloon pump was placed during heart transplant or for shock related to acute myocardial infarction. Invasive hemodynamics before and after intra-aortic balloon pump placement were assessed.

**Results:** We identified 23 patients with cardiac amyloid who had an intra-aortic balloon pump placed for heart failure-related cardiogenic shock. One-year survival was 74% and most (65%) were bridged to heart transplant while one was bridged to destination left ventricular assist device. Following intra-aortic balloon pump, mean arterial pressure, cardiac index, and cardiac power index were significantly increased, while mean right atrial pressure, mean pulmonary artery pressure, and pulmonary capillary wedge pressure were all significantly reduced. Smaller left ventricular end diastolic diameter (per cm) was associated with higher likelihood of a cardiac index of < 2.2 L/min/m^2^ following intra-aortic balloon pump (OR 0.16, CI 0.01 – 0.93, P=0.04).

**Conclusion:** Intra-aortic balloon pump significantly improved cardiac index while reducing right atrial pressure, mean pulmonary artery pressure, and pulmonary capillary wedge pressure in CA patients with heart failure-related cardiogenic shock.

## INTRODUCTION

The role of the intra-aortic balloon pump (IABP) as a form of temporary mechanical circulatory support (tMCS) has been well studied in acute myocardial infarction-related cardiogenic shock (AMI-CS) without a proven survival benefit. (1) Heart failure-related CS (HF-CS) represents distinct physiology from that of AMI-CS and previous studies suggest IABP use may improve hemodynamics and lead to clinical stabilization in this population. (2) Cardiac amyloidosis (CA) results in chronic heart failure (HF) from myocardial deposition of amyloid protein leading to restrictive physiology. The two most common types are transthyretin (ATTR) and light chain (AL)-CA. Despite promising therapeutic advances, such as Tafamidis for ATTR-CA and daratumumab-based treatments for AL-CA, many advance to stage D HF and may be considered for advanced therapies. (3, 4) Policy changes giving those with restrictive cardiomyopathy (RCM) United Network of Organ Sharing (UNOS) status 4 have led to higher heart transplantation (OHT) rates for CA. Additionally, those supported with IABP meeting defined hemodynamic criteria now receive UNOS status 2 classification, leading to higher rates of bridging to advanced therapies with IABP. (5) Few studies, however, have described IABP use in CA, perhaps from a lack of perceived hemodynamic benefit in this phenotype. It is also unclear if a role exists for IABP in bridging those with CA and HF-CS to advanced therapies.

Whereas bridging with left ventricular assist device (LVAD) is often limited by the smaller left ventricular (LV) dimensions and a higher propensity of bi-ventricular (Bi-V) failure due to LV and right ventricular (RV) involvement in CA, axillary IABP placement has made it feasible to support those with HF-CS in need of tMCS for longer periods while awaiting advanced therapies. (6, 7) The purpose of this study is severalfold: first, to describe the hemodynamic effects with IABP support in CA with HF-CS; second, to identify factors that might predict response to IABP; and lastly, to describe outcomes in CA with HF-CS who were supported with IABP.

## METHODS

### Study population

This study was approved by the Cleveland Clinic Institutional Review Board. Written informed consent was waived as all procedures were performed as part of routine clinical care. We retrospectively identified consecutive patients from our amyloid registry who had a diagnosis of either AL-CA or ATTR-CA and who were admitted to our institution’s intensive care unit (ICU) from January 2009 to January 2022. Patients were included in the study if they were 18 years or older, were admitted to our ICU with HF-CS, and, had an indwelling pulmonary artery catheter and IABP placed during the admission. Patients were excluded if IABP was placed during OHT or for AMI-CS. CS was defined hemodynamically as having the following present: Cardiac index (CI) of < 2.2 L/min/m^2^, a systolic blood pressure (SBP) < 90 mmHg, or the need for vasoactive medications and/or device intervention to maintain SBP > 90 mmHg. We retrospectively classified CS by The Society for Cardiovascular Angiography and Interventions (SCAI) CS Stage. SCAI stage C CS was classified as having the following present: hemodynamic instability (i.e., mean arterial pressure (MAP) < 60 mmHg or SBP < 90 mmHg), end-organ hypoperfusion (i.e., lactate ≥ 2 mmol/L, rise in creatinine ≥ 0.3 mg/dL from baseline), congestion (i.e., pulmonary capillary wedge pressure (PCWP) > 15 mmHg, elevated NT-pro BNP), and hemodynamic compromise (i.e., CI of < 2.2 L/min/m^2^). SCAI stage D CS was defined as a patient with stage C CS and with worsening CS (i.e., progressive end organ-hypoperfusion evidenced by rising lactate, lactate persistently >2 mmol/L, or worsening renal function) and/or worsening hemodynamics despite vasoactive medication and/or device support. (8) Indications for IABP placement were reviewed for each patient to confirm UNOS status 2 hemodynamic criteria was met for consideration for IABP support and required that the following be present in a 24-hour period, within 7 days of IABP placement: SBP < 90 mmHg, CI of < 1.8 L/min/m^2^ if not supported by vasoactive medication, or CI < 2.0 L/min/m^2^ if supported by vasoactive medication, and PCWP > 15 mmHg. (9)

Each patient in this study was identified from our CA registry and had a previously confirmed diagnosis of CA. Confirmation of diagnosis for ATTR-CA was by tissue biopsy with confirmation of ATTR deposits by mass spectrometry and/or immunohistochemistry staining. Alternatively, diagnosis using a non-biopsy approach was made following adherence to the American Society of Nuclear Cardiology (ASNC) published guidelines. (10, 11) Diagnosis of AL-CA was confirmed by histology showing light chain amyloid deposits with typing by immunohistochemistry or laser dissection mass spectrometry, or with a noncardiac biopsy in conjunction with echocardiography or cardiac magnetic resonance imaging suggestive of CA.

### Data collection

Demographic data was collected from the initial admission. Laboratory data was collected upon initial admission and 24 hours before and after IABP insertion. Serum creatinine was collected at 48-hours post-IABP. Patients with end-stage renal disease or on renal replacement therapy were excluded from comparison analysis. Total measured urine output (UOP) was collected within 24 hours before and after IABP insertion. Echocardiographic data and measurements were collected using the most recently available study before admission. The name and number of vasoactive medications were collected from admission and immediately before and after IABP insertion.

### Hemodynamic measurements and definitions

All non-invasive hemodynamic values including SBP, MAP, and heart rate (HR), were collected before, after, and at 48-hours post-IABP insertion. Invasive hemodynamic values were collected using the indwelling pulmonary artery catheter before, after, and at 48 hours post-IABP insertion and included the mean pulmonary artery pressure (mPAP), PCWP, mean RA pressure (RAP), CI (using estimated Fick method), pulmonary artery diastolic (PADP) and systolic pressure (PASP), systemic vascular resistance (SVR), and mixed venous oxygen saturation (MVO2). Calculated values included LV and RV cardiac power indices (CPI), calculated as [MAP x CI]/451 and [mPAP x CI]/451, respectively. Pulmonary artery pulsatility index (PAPi) was calculated as [PASP – PADP]/RAP, and SVR was calculated using the standard equation.

### Outcomes

Outcomes included 1-year survival from the index admission, upgrade to another form of MCS following IABP placement, and bridge to advanced therapy. Following IABP, indicators of response were considered based on changes in CI and PCWP. We defined lack of response to IABP as a CI remaining below 2.2 L/min/m^2^ and/or PCWP remaining above 20 mmHg.

### Statistical analyses

Continuous variables were expressed as mean values with associated standard deviation or median with associated interquartile range (IQR) for non-normally distributed variables. Categorical variables were expressed as numbers and percentages and were compared using Pearson Chi-squared test or Fisher exact test. For comparing hemodynamic variables before and after IABP insertion, Paired students t-test was used for normally distributed variables or Wilcoxon matched pairs test for non-normally distributed variables. Binary or linear regression analysis was used to determine if the following independent covariables were associated with response to IABP by our pre-defined criteria: mPAP, PCWP, CI, CPI, LVEDD, SVR, PAPi, LVEF, SBP, and creatinine. Statistical analyses were completed with Microsoft Excel (Microsoft, Redmond, WA) and GraphPad Prism version 9 (San Diego, California USA) software. A P-value of less than 0.05 was considered statistically significant.

## RESULTS

### Patient population

Between January 2001 and August 2021, a total of 1,563 patients received a diagnosis of CA. A total of 97 of these were admitted to our ICU between January 2009 and January 2022. Of these, 23 had hemodynamic evidence of HF-CS and had IABP placed; these patients formed our cohort. The age range for the cohort was 51 to 78 years. Underlying CA etiology was ATTR-CA in 52% and AL-CA in 48%. Upon admission, most patients were classified as SCAI stage C HF-CS. One patient had evidence of continued deterioration despite IABP insertion, and was classified as SCAI stage D HF-CS. All patients met hemodynamic criteria for UNOS status 2 for CS and consideration for IABP support (Table 1). Most patients were on a single vasoactive medication before and after IABP insertion (56%) and the majority were on the same vasoactive medication(s) post-IABP (70%) (Table 2).

**Table 1.** Baseline characteristics for CA patients with HF-CS and IABP.

| Parameter | All<br>N=23 |
| --- | --- |
| Demographics |  |
| Age, years | 68 [60 – 71] |
| Caucasian | 16 (70%) |
| Male | 18 (78%) |
| BMI 25-29, Kg/m<sup>2</sup> | 11 (48%) |
| BMI > 30, Kg/m<sup>2</sup> | 6 (26%) |
| Cardiac Amyloid Type |  |
| Transthyretin | 12 (52%) |
| Light Chain | 11 (48%) |
| Comorbidities |  |
| Stroke | 6 (26%) |
| Diabetes | 5 (22%) |
| CAD | 6 (26%) |
| SCAI CS Stage |  |
| Stage C | 22 (96%) |
| Stage D | 1 (4%) |
| Laboratory |  |
| Troponin T, ng/L | 1.07 ±1.78 |
| NT pro BNP, pg/mL | 17,437 [3,862 – 30,990] |
| Creatinine, mg/dL | 2.15 ±0.98 |
| Albumin, g/dL | 3.4 ±0.3 |
| Serum lactate, mmol/L | 2.47 ±2.3 |
| Serum bicarbonate, mmol/L | 25 ±4 |
| Echocardiographic |  |
| LVEF, % | 37 ±14 |
| LVEDD, cm | 4.0 ±0.5 |
| LV posterior wall thickness, cm | 1.67 ±0.4 |
| IVS wall thickness, cm | 1.72 ±0.5 |
| Mitral Regurgitation (≥ 3+) | 2 (9%) |
| Tricuspid Regurgitation (≥ 3+) | 5 (22%) |

**Abbreviations:** IABP = intra-aortic balloon pump, SCAI = The Society for Cardiovascular Angiography and Interventions, CS = cardiogenic shock, CA = cardiac amyloid, LVEF = left ventricular ejection fraction, LVEDD = left ventricular end diastolic diameter, NT pro BNP = N-Terminal Pro B-type Natriuretic Peptide, BMI = body mass index, IVS = interventricular septum.

**Table 2.**
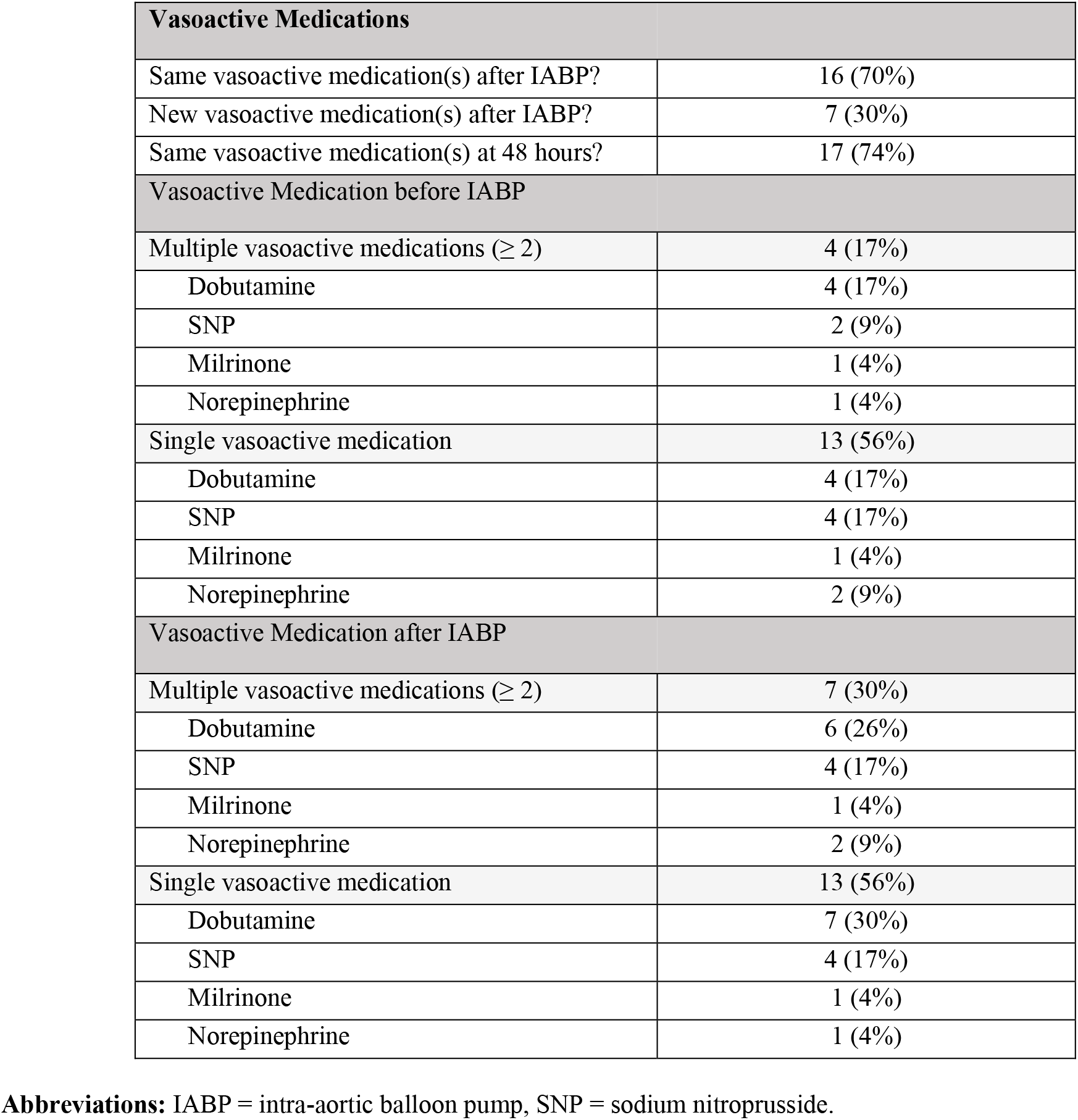
Vasoactive medications before and after IABP insertion.

| **Vasoactive Medications** |  |
| --- | --- |
| Same vasoactive medication(s) after IABP? | 16 (70%) |
| New vasoactive medication(s) after IABP? | 7 (30%) |
| Same vasoactive medication(s) at 48 hours? | 17 (74%) |
| **Vasoactive Medication before IABP** |  |
| Multiple vasoactive medications ($\geq 2$) | 4 (17%) |
| Dobutamine | 4 (17%) |
| SNP | 2 (9%) |
| Milrinone | 1 (4%) |
| Norepinephrine | 1 (4%) |
| Single vasoactive medication | 13 (56%) |
| Dobutamine | 4 (17%) |
| SNP | 4 (17%) |
| Milrinone | 1 (4%) |
| Norepinephrine | 2 (9%) |
| **Vasoactive Medication after IABP** |  |
| Multiple vasoactive medications ($\geq 2$) | 7 (30%) |
| Dobutamine | 6 (26%) |
| SNP | 4 (17%) |
| Milrinone | 1 (4%) |
| Norepinephrine | 2 (9%) |
| Single vasoactive medication | 13 (56%) |
| Dobutamine | 7 (30%) |
| SNP | 4 (17%) |
| Milrinone | 1 (4%) |
| Norepinephrine | 1 (4%) |

**Abbreviations:** IABP = intra-aortic balloon pump, SNP = sodium nitroprusside.

### Baseline labs and hemodynamics

Mean lactate prior to IABP was 2.47 ±2.3 mmol/L and serum creatinine prior to IABP insertion was 2.15 ±0.98 mg/dL. Median NT pro BNP was significantly elevated at 17,437 [IQR 3,862 – 30,990] pg/mL. Mean LVEDD was 4.0 ±0.5 cm and mean left ventricular ejection fraction (LVEF) was 37 ±14 percent. All hemodynamic measurements were collected within 24 hours before and after IABP insertion, with median time of assessment being 5 [IQR 3 – 9] and 10 [6 – 14] hours, respectively. The entire cohort had a CI below 2.0 L/min/m^2^ upon admission to the ICU, and nearly all patients had a CI < 2.2 L/min/m^2^ within the 24 hours prior to IABP insertion. Initial PCWP was ≥ 20 mmHg in 87% of the cohort. Initial RAP and mPAP were also significantly elevated (Table 3).

**Table 3.** Hemodynamics Before and After IABP.

| Parameter | Before IABP | After IABP | Mean Change | P value |
| --- | --- | --- | --- | --- |
| Mean arterial pressure, mmHg | 67.4±9.8 | 74.7±11.7 | 7.2±13.9 | 0.02 |
| RAP, mmHg | 18.6±7.9 | 14.3±5.2 | -4.2±7.3 | 0.01 |
| PA diastolic pressure, mmHg | 26.4±6.1 | 22.1±6.2 | -4.3±4.9 | 0.0004 |
| PA systolic pressure, mmHg | 51.2±11.9 | 45.7±12.9 | -5.5±9.1 | 0.008 |
| Mean PA pressure, mmHg | 34.6±6.9 | 30.5±7.1 | -4.0±5.9 | 0.003 |
| PCWP, mmHg | 25±6 | 20±5.4 | -4.0±4.9 | 0.001 |
| Mixed Venous O2 saturation | 47.3±10.3 | 56.7±8.3 | 9.3±9.5 | 0.0001 |
| Cardiac Index, liters/min/m<sup>2</sup> | 1.67±0.37 | 2.17±0.57 | 0.49±0.46 | <0.0001 |
| LV Cardiac Power Index, W/m<sup>2</sup> | 0.25±0.06 | 0.35±0.1 | 0.10±0.10 | <0.0001 |
| RV Cardiac Power Index, W/m<sup>2</sup> | 0.12±0.04 | 0.145±0.05 | 0.016±0.03 | 0.04 |
| PA Pulsatility Index | 1.7±1.2 | 2.06±1.5 | 0.335±1 | 0.15 |
| RAP/PCWP | 0.77±0.30 | 0.73±0.23 | -0.03±0.35 | 0.34 |
| Systemic vascular resistance, dyne/s/cm | 1390±396 | 1210±377 | -185±261 | 0.004 |

**Abbreviations:** RAP = mean right atrial pressure, PA = pulmonary artery, PCWP = pulmonary capillary wedge pressure, IABP = intra-aortic balloon pump, LV = left ventricular, RV = right ventricular.

### Labs and hemodynamics after IABP

Following IABP, MAP was significantly increased, and hemodynamic function improved significantly, evidenced by increased CI and LV CPI. RV CPI also significantly increased following IABP (Figure 1, Table 3). Of the 22 patients who had a CI below 2.2 L/min/m^2^ before IABP, 50% improved to a CI of ≥ 2.2 L/min/m^2^ immediately after IABP. Mean PCWP was significantly lower following IABP and 52% achieved a PCWP < 20 mmHg. RAP and mPAP were also significantly reduced following IABP (Figure 1, Table 3). At 48 hours post-IABP insertion, CI was 2.49±0.7, and 73% had a CI of ≥ 2.0 L/min/m^2^. Compared to the initial post-IABP hemodynamics, PCWP, RAP, and mPAP at 48 hours post-IABP were not significantly different (Figure 2). Serum lactate following IABP was significantly lower (1.7 ±0.65 vs 2.47 ±2.3, P=0.01). Serum creatinine post-IABP insertion was not significantly different from baseline (2.10 ±1.1 vs 2.15 ±0.98, P=0.60). Total measured UOP before and after IABP was not significantly different (Figure 3).

**Figure 1.**
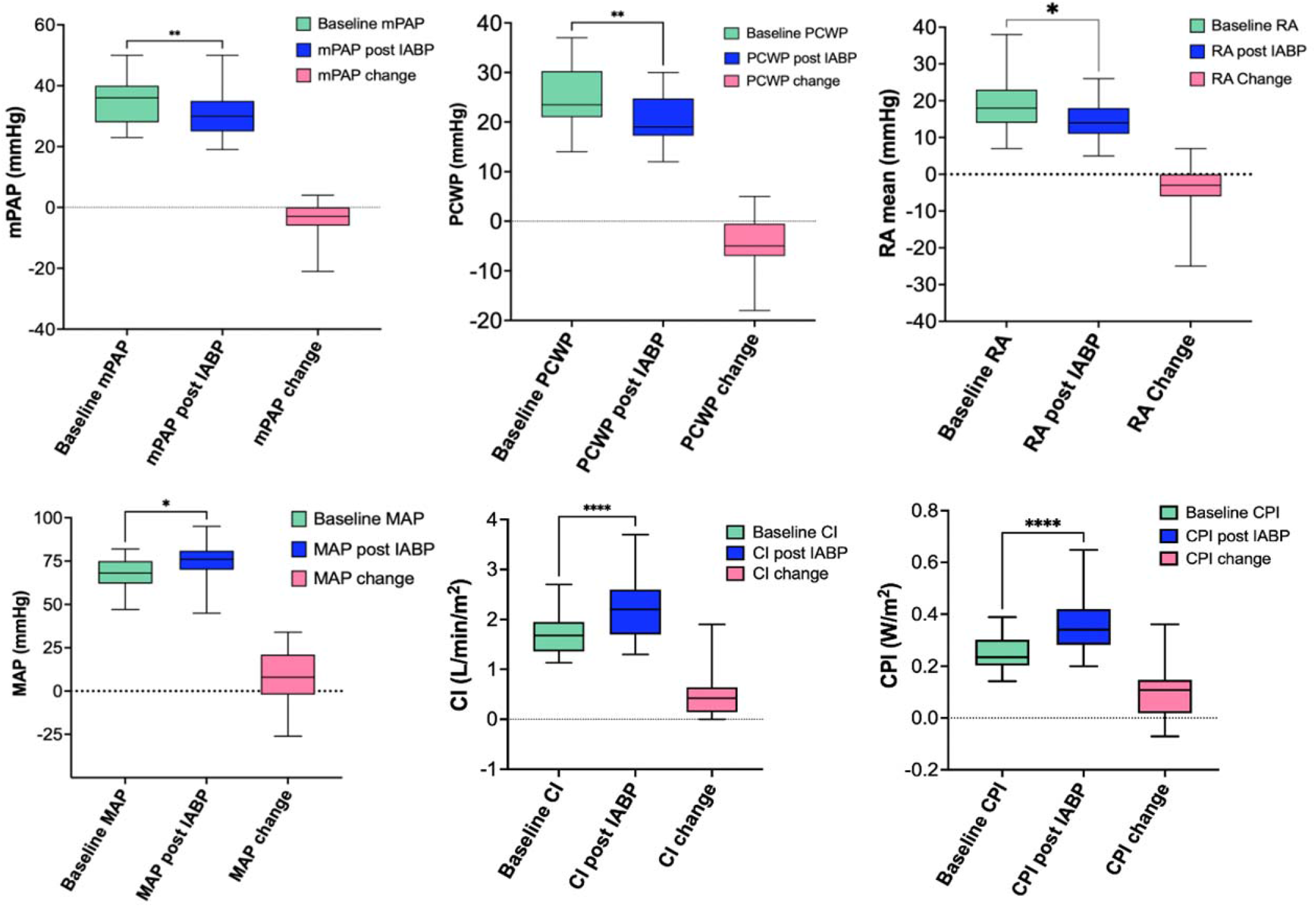
Hemodynamic Parameters Before and After IABP. **Abbreviations:** mPAP = mean pulmonary artery pressure, RA= right atrial pressure, PCWP = pulmonary capillary wedge pressure, MAP = mean arterial pressure, CPI = cardiac power index, CI = cardiac index, IABP = intra-aortic balloon pump.

**Figure 2.**
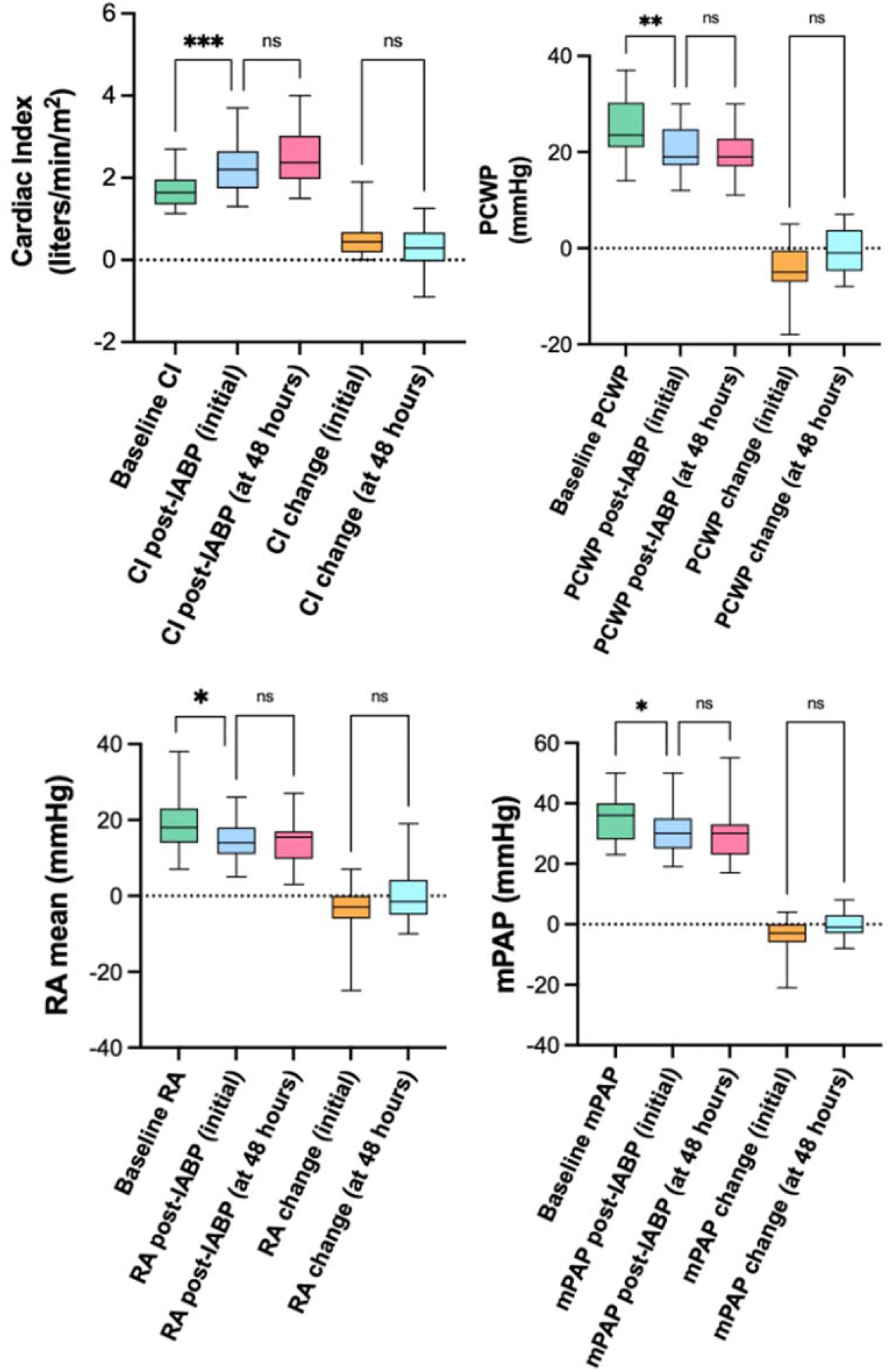
Hemodynamic changes at 48-hours post-IABP. **Abbreviations:** IABP = intra-aortic balloon pump, CI = cardiac index, RA = mean right atrial pressure, PCWP = pulmonary capillary wedge pressure, mPAP = mean pulmonary artery pressure.

**Figure 3.**
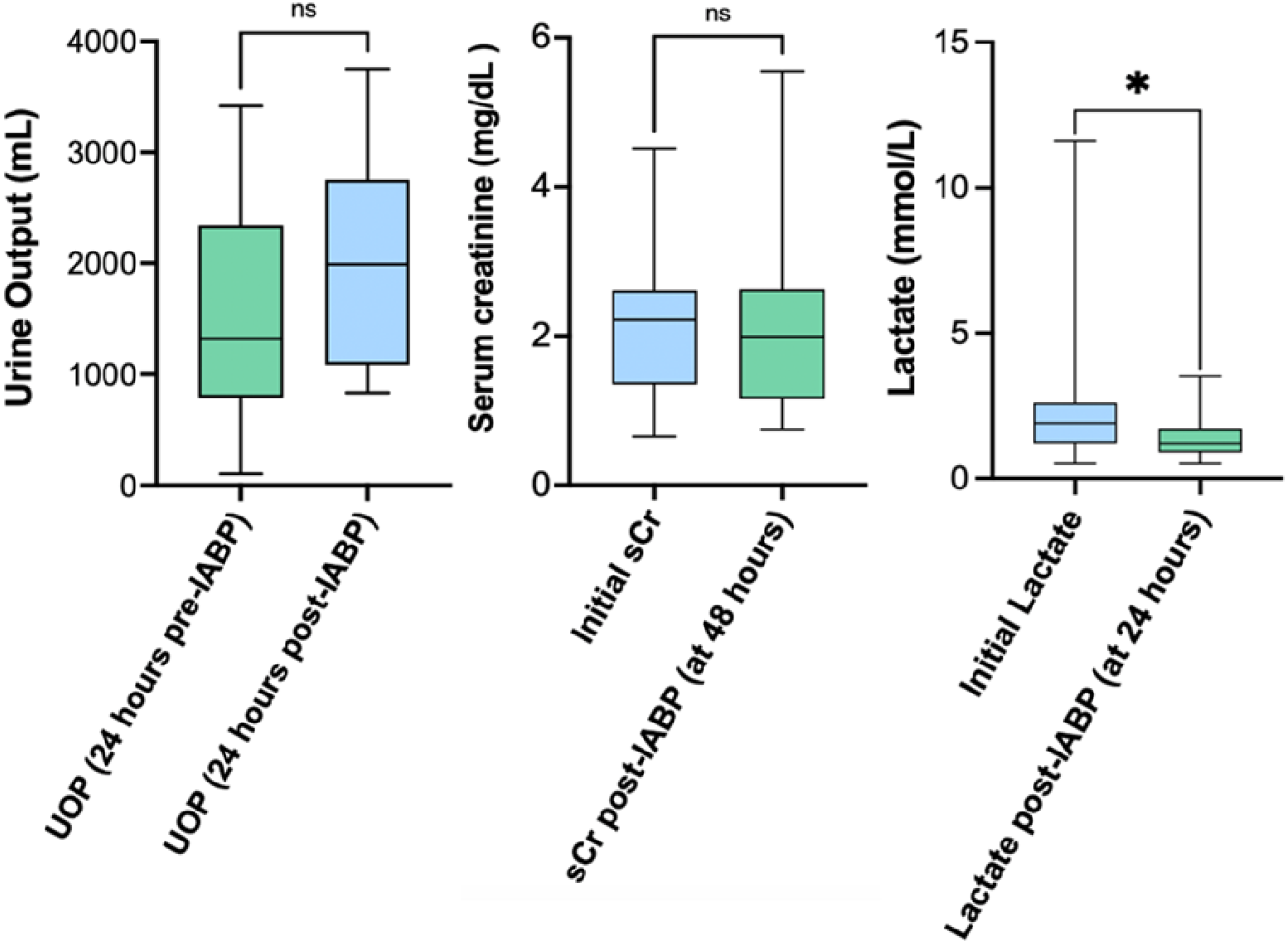
Measuring change in UOP, lactate, and creatinine before and after IABP. **Abbreviations:** UOP = urine output. IABP = intra-aortic balloon pump, sCr = serum creatinine.

### Predictors of response to IABP

Response to IABP was assessed using available hemodynamics before and after IABP. To follow our pre-defined criteria for response, the single patient with a CI of > 2.2 L/min/m^2^ before IABP was excluded from analysis for predictors of response by CI. Univariable logistic regression revealed that LVEDD size was associated with CI following IABP, specifically, smaller LVEDD size (per cm) was associated with higher likelihood of having a CI of < 2.2 L/min/m^2^ following IABP (OR 0.16, CI 0.01 – 0.93, P=0.04). A similar relationship between LVEDD size (per cm) and CI was noted using a simple linear regression model, and was statistically significant (R^2^ = 0.23, CI [0.065 – 0.850], p=0.02), revealing larger LVEDD size (per cm) predicted higher CI after IABP. Higher pre-IABP SVR was associated with a significantly higher likelihood of a CI of < 2.2 L/min/m^2^ following IABP (Table 3). Higher serum creatinine before IABP insertion was significantly associated with a PCWP of > 20 mmHg after IABP (Table 4).

**Table 4.** Predicting Response to IABP in CA.

| **Predicting response to IABP** |  |  |  |
| --- | --- | --- | --- |
| **Univariable Regression analysis** |  |  |  |
|  | Odds Ratio | 95% CI | P-value |
| **Cardiac Index < 2.2 L/min/m<sup>2</sup> after IABP** |  |  |  |
| SVR, dyne/s/cm | 1.004 | 1.001 – 1.008 | 0.008 |
| LVEDD, cm | 0.16 | 0.015 – 0.935 | 0.04 |
| Initial SBP, mmHg | 1.01 | 0.925 – 1.116 | 0.77 |
| Pulmonary artery pulsatility index | 1.08 | 0.4564 – 2.634 | 0.85 |
| LVEF, % | 1.00 | 0.946 – 1.067 | 0.99 |
| **PCWP > 20 mmHg after IABP** |  |  |  |
| Creatinine, mg/dL | 3.8 | 1.226 – 22.26 | 0.007 |
| RAP, mmHg | 1.06 | 0.9456 – 1.232 | 0.29 |
| Pulmonary artery pulsatility index | 0.66 | 0.2204 – 1.607 | 0.38 |
| Log NT pro BNP, pg/L | 3.8 | 0.667 – 34.85 | 0.13 |
| LVEF, % | 1.02 | 0.9611 – 1.093 | 0.50 |

**Abbreviations:** IABP = intra-aortic balloon pump, SVR = systemic vascular resistance, SBP = systolic blood pressure, LVEF = left ventricular ejection fraction, RAP = mean right atrial pressure, MAP = mean arterial pressure, mPAP = mean pulmonary artery pressure, PCWP = pulmonary capillary wedge pressure, CI = cardiac index, NT pro BNP = N-Terminal B-type Pro-Natriuretic Peptide.

### Patient outcomes

Overall 1-year survival was 74% (N=17). Of those who survived, 65% (N=15) were successfully bridged to OHT and one was bridged to destination therapy LVAD. Of those bridged to advanced therapies, 75% (N=12) were stabilized and bridged with IABP alone. Four patients required tMCS upgrade to extra-corporeal membrane oxygenation, 2 due to worsening CS and 2 due to pulseless electrical activity arrest. The overall time to IABP insertion was 1 [IQR 4 to 16] days. Those bridged to advanced therapies were supported with IABP for 9 [IQR 6.25 to 17.25] days. Time to advanced therapies from IABP insertion was 13 [IQR 7.25 to 23.75] days. For those not bridged, median time to IABP insertion was 2 [IQR 0 to 16] days and duration of IABP support was 4 [IQR 3 to 14] days. Overall hospital duration for those bridged to advanced therapies was 53 [IQR 41 to 75.5] days and was 19 [IQR 5 to 30] days for those not bridged. Three patients were discharged home with hospice without further support and 3 experienced in-hospital mortality despite IABP. (Table 5).

**Table 5.** Outcomes in Patients with CA and HF-CS supported by IABP.

| **Outcomes** |  |
| --- | --- |
| Entire Cohort | N=23 |
| Length of IABP placement, days | 9 [4 – 15] |
| Survival at 1 year, % | 17 (74%) |
| Bridged to advanced therapies | N=16 (70%) |
| OHT, % | 15 (65%) |
| LVAD, % | 1 (4%) |
| No device upgrade, % | 12 (75%) |
| Required device upgrade, % | 4 (25%) |
| Time to OHT (from IABP insertion), days | 13 [7.25 – 23.75] |
| Hospitalization duration (OHT/LVAD), days | 53 [41 to 75.5] |
| Not bridged to advanced therapies | N=7 (30%) |
| Hospitalization duration (not bridged), days | 19 [5 to 30] |
| Discharged with Hospice, % | 3 (13%) |
| In hospital mortality, % | 3 (13%) |

**Abbreviations:** IABP = intra-aortic balloon pump, LVAD = left ventricular assist device, OHT = open heart transplant.

## DISCUSSION

In this study we provide an in-depth analysis of the hemodynamic response with IABP in patients with CA and HF-CS. Our primary findings are first, in those with CA and HF-CS, there was significant augmentation of CI following IABP and significant reductions in LV and RV filling pressures. Second, predictors of response to IABP by CI were a smaller baseline LVEDD size per cm and higher pre-IABP SVR. Lastly, of those bridged to advanced therapies, most were successfully stabilized and bridged with IABP alone, suggesting that IABP may be a feasible modality for bridging those with CA and SCAI stage C HF-CS to advanced therapies.

CS is a low output state characterized by end-organ hypoperfusion. Vasoactive medications are often used in the initial management of CS and IABP is a commonly utilized tMCS device. The decision to utilize the IABP depends on the etiology of CS, the associated complications, and severity of CS at presentation. (12) A different hemodynamic response to IABP might be expected when CS occurs in chronic HF as the result of decompensation from a chronic low-output state, compared with CS from coronary ischemia in which myocardial contractility is acutely impaired. Retrospective studies comparing IABP use in AMI-CS and HF-CS have shown a greater impact on hemodynamics in HF-CS, specifically, the augmentation of cardiac output. (13-14) This emphasizes the importance of the etiology of CS and highlights differences in underlying physiology. Prior retrospective studies investigating the use of IABP in HF-CS provide insights as to a role for IABP in stabilization and possibly as a bridge to advanced therapies. (15) Our study, however, provides important insights into the hemodynamic response to IABP counter-pulsation in those with CA and HF-CS. Specifically, we describe significant improvements in both CI and LV and RV CPI following IABP insertion. The hemodynamic findings of our study are in line with prior data; despite the unique underlying restrictive physiology, those with CA and HF-CS demonstrated benefit from LV unloading with IABP, evidenced by the significant improvement in hemodynamic function.

The utility of vasoactive drugs that reduce afterload in acute HF, such as SNP, has been demonstrated. (16) During the deflation phase of the IABP, LV afterload is reduced leading to reduced LV wall stress and myocardial oxygen consumption. We describe a relationship between LVEDD and CI with IABP, specifically, that smaller LVEDD size per cm was associated with a higher likelihood of a CI < 2.2 L/min/m^2^ following IABP unloading. One prior study demonstrated a relationship between larger LVEDD size and response to SNP in acute HF. (17) One possible explanation, based on the Frank Starling curve, is that responsiveness to afterload reduction might be more pronounced at the higher LV filling pressures (preload) that accompany larger LVEDD’s. However, this has not been previously described in the context of CA. The observation that those with CA and HF-CS in our cohort were able to significantly augment their CI in response to afterload reduction with IABP therapy is a positive finding. The predictive value of LVEDD in the response to afterload reduction with IABP and SNP should be further investigated in this population.

UNOS policy changes in 2018 giving those with IABP status 2 designation have led to increased rates of bridging with IABP and improved waitlist mortality. Post-OHT outcomes for those bridged with IABP have been, for the most part, comparable. (18) Our study provides valuable insights for the potential of IABP as a beneficial and feasible bridging modality to advanced therapies in CA, specifically, for those patients who present in SCAI stage C HF-CS, and/or for those in whom durable or tMCS might be prohibitive. Of those successfully bridged to advanced therapies in our cohort, the majority (75%) did not require tMCS upgrade and were stabilized with IABP alone. Furthermore, regarding post-OHT or LVAD outcomes, all patients who were bridged to advanced therapies in our study were alive at 1 year following the index ICU admission. Literature examining tMCS in CA with CS is limited. Successful bridging with durable LVAD is feasible, yet highly dependent on patient selection, and is often restricted by smaller LV cavity dimensions and concomitant RV failure. (19) Durable Bi-V support (i.e., BiVAD, total artificial heart) has shown some promise in bridging to advanced therapies in CA in retrospective analyses. (20-22) Patients in these studies, however, were generally sicker (INTERMACS 1 and 2), whereas most in our study had SCAI stage C HF-CS. Compared to LVAD, which requires intrinsic RV function to maintain adequate LV preload, IABP is not dependent on RV function which is often impaired in CA. Furthermore, we describe significant improvement in RV CPI following IABP, possibly owing to improved right to left-sided flow with LV unloading. Prior studies have demonstrated improved RV function with IABP, however, not in the CA population. (23) Compared with LVAD, IABP placement is not precluded by LV cavity size and percutaneous placement of IABP in the axillary position makes extended support more feasible. Further prospective studies are warranted to validate the utility of IABP for bridging CA patients with HF-CS to advanced therapies, specifically those who can be stabilized with IABP alone.

### Limitations

Our study was a single-center study using registry data from our large quaternary referral center, which might have introduced referral bias. Additionally, most patients were classified as having SCAI stage C HF-CS, limiting the generalizability of our study. Selecting patients admitted to our ICU, who had a pulmonary artery catheter and IABP placed may have introduced selection bias. Our relatively small sample size further limits generalizability and may have led to an inability to detect significant differences within the cohort.

## CONCLUSION

When utilized in the context of CA with HF-CS, IABP therapy significantly improved CI. Augmentation of CI with IABP was less effective in those with smaller LVEDD size. Successful bridging to advanced therapies with excellent 1-year survival rates suggest that IABP may be a useful modality to bridge those with CA and SCAI stage C HF-CS to advanced therapies.

## Data Availability

The data, analytic methods, and study materials will not be made available to other researchers for purposes of reproducing the results or replicating the procedures. The authors had full access to all the data in the study and take responsibility for the integrity of the data and accuracy of the data analysis, and may agree to make data available upon reasonable request.

## NON-STANDARD ABBREVIATIONS AND ACRONYMS

CA: cardiac amyloidosis
IABP: intra-aortic balloon pump
HF: heart failure
MCS: mechanical circulatory support
ATTR-CA: transthyretin cardiac amyloidosis
AL-CA: light chain cardiac amyloidosis
CI: cardiac index
CPI: cardiac power index
PCWP: pulmonary capillary wedge pressure
RAP: right atrial pressure
mPAP: mean pulmonary artery pressure
HF: heart failure
CS: cardiogenic shock

## Author Contributions

Drs Longinow and Lee designed the research study. Dr. Longinow analyzed the data and wrote the initial draft of the manuscript. All authors provided critical input and revisions for the manuscript.

## REFERENCES

1. Thiele H, Zeymer U, Neumann FJ, Ferenc M, Olbrich HG, Hausleiter J, Richardt G, Hennersdorf M, Empen K, Fuernau G, Desch S, Eitel I, Hambrecht R, Fuhrmann J, Böhm M, Ebelt H, Schneider S, Schuler G, Werdan K; IABP-SHOCK II Trial Investigators. Intraaortic balloon support for myocardial infarction with cardiogenic shock. N Engl J Med. 2012 Oct 4;367(14):1287–96.

2. Sintek MA, Gdowski M, Lindman BR, Nassif M, Lavine KJ, Novak E, Bach RG, Silvestry SC, Mann DL, Joseph SM. Intra-Aortic Balloon Counterpulsation in Patients With Chronic Heart Failure and Cardiogenic Shock: Clinical Response and Predictors of Stabilization. J Card Fail. 2015 Nov;21(11):868–876.

3. Maurer, M. S., Schwartz, J. H., Gundapaneni, B., Elliott, P. M., Merlini, G., Waddington-Cruz, M., Kristen, A. V., Grogan, M., Witteles, R., Damy, T., Drachman, B. M., Shah, S. J., Hanna, M., Judge, D. P., Barsdorf, A. I., Huber, P., Patterson, T. A., Riley, S., Schumacher, J., Stewart, M., … ATTR-ACT Study Investigators (2018). Tafamidis Treatment for Patients with Transthyretin Amyloid Cardiomyopathy. The New England journal of medicine, 379(11), 1007–1016.

4. Witteles RM. Cardiac Transplantation and Mechanical Circulatory Support in Amyloidosis. JACC CardioOncol. 2021;3(4):516–521. Published 2021 Oct 19.

5. Griffin JM, Chiu L, Axsom KM, et al. United Network for Organ Sharing outcomes after heart transplantation for AL compared to ATTR cardiac amyloidosis. Clin Transplant. 2020;34: e14028.

6. Grupper A, Park SJ, Pereira NL, et al. Role of ventricular assist therapy for patients with heart failure and restrictive physiology: Improving outcomes for a lethal disease. J Heart Lung Transplant. 2015;34(8):1042–1049.

7. Bhimaraj, A., Agrawal, T., Duran, A., Tamimi, O., Amione-Guerra, J., Trachtenberg, B., Guha, A., Hussain, I., Kim, J., Kassi, M., Xu, J., Suarez, E., Ngo, U. Q., Torre-Amione, G., & Estep, J. D. (2020). Percutaneous Left Axillary Artery Placement of Intra-Aortic Balloon Pump in Advanced Heart Failure Patients. JACC. Heart failure, 8(4), 313–323.

8. Naidu S, Baran D, Jentzer J, et al. SCAI SHOCK Stage Classification Expert Consensus Update: A Review and Incorporation of Validation Studies. J Am Coll Cardiol. 2022 Mar, 79 (9) 933–946.

9. U.S. Department of Health and Human Services. Organ Procurement and Transplantation Network: adult heart allocation. Available at: https://optn.transplant.hrsa.gov/learn/professional-education/adult-heart-allocation/.

10. Hanna M, Ruberg FL, Maurer MS, Dispenzieri A, Dorbala S, Falk RH, Hoffman J, Jaber W, Soman P, Witteles RM, Grogan M. Cardiac Scintigraphy With Technetium-99m-Labeled Bone-Seeking Tracers for Suspected Amyloidosis: JACC Review Topic of the Week. Journal of American College of Cardiology. 2020 Jun 9;75(22):2851–2862.

11. Dorbala S, Ando Y, Bokhari S, Dispenzieri A, Falk RH, Ferrari VA, Fontana M, Gheysens O, Gillmore JD, Glaudemans AWJM, Hanna MA, Hazenberg BPC, Kristen AV, Kwong RY, Maurer MS, Merlini G, Miller EJ, Moon JC, Murthy VL, Quarta CC, Rapezzi C, Ruberg FL, Shah SJ, Slart RHJA, Verberne HJ, Bourque JM. ASNC/AHA/ASE/EANM/HFSA/ISA/SCMR/SNMMI Expert Consensus Recommendations for Multimodality Imaging in Cardiac Amyloidosis: Part 1 of 2-Evidence Base and Standardized Methods of Imaging. J Card Fail. 2019 Nov;25(11):e1–e39. doi: 10.1016/j.cardfail.2019.08.001. Epub 2019 Aug 29. Erratum in: J Card Fail. 2022 Jul;28(7):e1-e4. PMID: 31473268.

12. van Diepen S, Katz JN, Albert NM, Henry TD, Jacobs AK, Kapur NK, Kilic A, Menon V, Ohman EM, Sweitzer NK, Thiele H, Washam JB, Cohen MG; American Heart Association Council on Clinical Cardiology; Council on Cardiovascular and Stroke Nursing; Council on Quality of Care and Outcomes Research; and Mission: Lifeline. Contemporary Management of Cardiogenic Shock: A Scientific Statement From the American Heart Association. Circulation. 2017 Oct 17;136(16):e232–e268.

13. Prondzinsky Roland* Unverzagt Susanne† Russ, Martin Lemm*, Henning Swyter*, Michael Wegener*, Nikolas Buerke*, Ute Raaz*, Uwe Ebelt*, Henning Schlitt*, Axel Heinroth*, Konstantin Haerting*, Johannes Werdan†, Karl Buerke Michael**. Hemodynamic Effects of Intra-aortic Balloon Counterpulsation in Patients With Acute Myocardial Infarction Complicated by Cardiogenic Shock: The Prospective, Randomized IABP Shock Trial. Shock: April 2012 - Volume 37 - Issue 4 - p 378–384

14. Malick W, Fried JA, Masoumi A, Nair A, Zuver A, Huang A, Haythe J, Farr M, Rabbani L, Karmpaliotis D, Kirtane AJ, Topkara VK, Takeda K, Garan AR. Comparison of the Hemodynamic Response to Intra-Aortic Balloon Counterpulsation in Patients With Cardiogenic Shock Resulting from Acute Myocardial Infarction Versus Acute Decompensated Heart Failure. Am J Cardiol. 2019 Dec 15;124(12):1947–1953.

15. Brown MA, Sheikh FH, Ahmed S, Najjar SS, Molina EJ. Intra-Aortic Balloon Pump as a Bridge to Durable Left Ventricular Assist Device. J Am Heart Assoc. 2021;10(15):e019376.

16. Mullens W, Abrahams Z, Francis GS, Skouri HN, Starling RC, Young JB, Taylor DO, Tang WH. Sodium nitroprusside for advanced low-output heart failure. J Am Coll Cardiol. 2008 Jul 15;52(3):200–207.

17. Garatti L, Frea S, Bocchino PP, Angelini F, Cingolani M, Sacco A, Rondinara GM, Bagnardi V, Sala IM, Kapur NK, Colombo PC, De Ferrari GM, Morici N. Sodium nitroprusside in acute heart failure: A multicenter historic cohort study. Int J Cardiol. 2022 Dec 15;369:37–44.

18. Huckaby, L. V., Seese, L. M., Mathier, M. A., Hickey, G. W., & Kilic, A. (2020). Intra-Aortic Balloon Pump Bridging to Heart Transplantation: Impact of the 2018 Allocation Change. Circulation. Heart failure, 13(8), e006971.

19. Randhawa VK, Gabrovsek A, Soltesz EG, et al. A case series of cardiac amyloidosis patients supported by continuous-flow left ventricular assist device. ESC Heart Fail. 2021;8(5):4353–4356.

20. Chen Q, Moriguchi J, Levine R, et al. Outcomes of Heart Transplantation in Cardiac Amyloidosis Patients: A Single Center Experience. Transplant Proc. 2021;53(1):329–334.

21. Kittleson MM, Cole RM, Patel J, et al. Mechanical circulatory support for cardiac amyloidosis. Clin Transplant. 2019;33:e13663.

22. Michelis KC, Zhong L, Tang WHW, et al. Durable Mechanical Circulatory Support in Patients With Amyloid Cardiomyopathy: Insights From INTERMACS. Circ Heart Fail. 2020;13(12):e007931.

23. Liakopoulos, O. J., Ho, J. K., Yezbick, A. B., Sanchez, E., Singh, V., & Mahajan, A. (2010). Right ventricular failure resulting from pressure overload: role of intra-aortic balloon counterpulsation and vasopressor therapy. The Journal of surgical research, 164(1), 58–66.

